# An Explainable Multimodal AI Framework with Reinforcement Learning for Post-Surgical Clinical Decision Support

**DOI:** 10.64898/2026.06.08.26355217

**Authors:** Mostak Ahmed, Faisal Ahmed, Shirajum Munira Mow, Papeya Akter Taha, Sanchayan Barua, Md Mostafizur Rahman, Al Rafy, Sadnan Mohosin Mondol, Mohammad Imtiaz Faisal

## Abstract

Post-surgical adverse outcomes, including mortality, intensive care readmission, and complications, remain major challenges for clinical decision-making. Existing machine learning approaches focus on outcome prediction while operating as opaque systems, limiting clinical trust and the translation of predictions into treatment decisions, and many clinical studies rely on synthetic data in which shared intermediate variables create circular dependencies between inputs and targets that compromise reported performance. We aimed to develop an explainable multimodal architecture and a rigorous evaluation methodology that address these gaps. We designed a two-stage architecture integrating supervised deep learning for risk prediction with conservative Q-learning for action recommendation. The first stage uses five modality-specific encoders for structured records, physiological time-series, chest radiographs, clinical notes, and surgical metadata, unified through cross-modal attention into a shared patient-state representation. The second stage applies offline reinforcement learning to recommend clinical actions while preventing value overestimation. We formally characterized a target-leakage flaw in synthetic pipelines and propose a real-data methodology using a verified clinical database, with event-censored temporal separation and uncertainty-weighted per-task training. Component-level behavior was validated on a controlled synthetic benchmark, demonstrating that the architecture functions as designed without claiming clinical validity. The cross-modal attention and risk-prediction components behaved as expected, whereas the offline reinforcement learning stage did not converge on the benchmark, indicating that value estimation requires further investigation on real clinical data. The architecture provides dual-level explainability through attention visualization and value decomposition, contributing a deployable design, a formal methodological critique of synthetic-data practices, and a complete framework for clinically valid evaluation.

## 1. Introduction

Post-surgical complications remain a leading cause of morbidity and mortality in hospitalized patients, with adverse outcomes including unexpected intensive care unit (ICU) admissions, major complications, and death imposing substantial burdens on patients, healthcare systems, and society. A recent global analysis estimated that 4.2 million people die within 30 days of surgery each year, making postoperative death the third leading cause of death globally after ischemic heart disease and stroke [1]. The global volume of surgery—estimated at over 310 million procedures annually [2]—underscores both the scale of the problem and the potential impact of improved risk stratification systems. Surgical adverse events remain common even in high-resource settings, with studies documenting adverse event rates between 3% and 17% across different surgical specialties [3].

Early identification of high-risk patients and timely intervention can significantly improve outcomes, yet the complexity of perioperative care—involving heterogeneous data sources, time-sensitive decisions, and patient-specific factors—makes consistent risk assessment challenging even for experienced clinicians. Traditional scoring systems such as APACHE and SOFA provide useful but limited risk stratification, typically relying on small numbers of manually collected variables and lacking the capacity to integrate the full spectrum of available clinical data.

Machine learning approaches to surgical outcome prediction have proliferated in recent years, with deep learning demonstrating the ability to capture complex non-linear relationships among clinical variables that traditional scoring systems may miss [4,5]. However, three fundamental limitations persist in the current landscape of clinical AI for surgical care.

First, most systems function as prediction-only tools that estimate what might happen without providing actionable guidance on what should be done, leaving clinicians to independently translate probabilistic risk scores into concrete treatment decisions [6]. This gap between prediction and action represents a major barrier to clinical impact. Second, the predominance of black-box models limits clinical adoption, as practitioners require understanding of the reasoning behind predictions to integrate AI recommendations safely into their workflow [7,8]. Without interpretability, even accurate models face resistance from clinical stakeholders who bear responsibility for patient outcomes.

Third—and perhaps most critically—many clinical AI studies rely on synthetic or semi-synthetic datasets where both input features and outcome labels are derived from the same underlying generative process, creating circular dependencies that inflate performance metrics without testing genuine clinical prediction capability [9,10]. This methodological concern is particularly acute in multimodal frameworks, where the complexity of multi-step data generation pipelines can obscure the relationship between inputs and targets, making it difficult to distinguish genuine learning from mathematical artifact recovery.

This work presents a two-stage explainable multimodal architecture for post-surgical clinical decision support, with explicit attention to methodological rigor in the design of both the model architecture and the evaluation pipeline. Our contributions are:

1. **Identification and Characterization of Target Leakage:** We formally document how intermediate risk scores used to simultaneously generate input features and outcome labels in synthetic clinical data pipelines create closed mathematical loops that render resulting performance metrics scientifically invalid, and we demonstrate this phenomenon empirically.
2. **Real-Data Evaluation Methodology:** We specify a complete pipeline for extracting verified prediction targets and unmodulated multimodal inputs from MIMIC-IV, with event-censored temporal separation that closes the input window at the moment of the first outcome event, preventing the temporal information leakage present in fixed-window designs.
3. **Multimodal Fusion Architecture:** We develop modality-specific encoders for five clinical data types—structured EHR, physiological time-series, chest radiographs, clinical notes, and surgical metadata—unified through cross-modal attention into a shared 512-dimensional patient state representation suitable for both prediction and decision support.
4. **Two-Stage Prediction-to-Action Pipeline with Dual-Level Explainability:** Stage-1 provides probabilistic risk estimates using Focal Loss with uncertainty-weighted per-task training; Stage-2 uses Conservative Q-Learning with convergence-gated deployment for action recommendation. Both stages support interpretability through attention visualization, feature attribution, and Q-value decomposition.

We validate the architecture’s component-level behavior on a controlled synthetic benchmark— explicitly framed as testing architectural function rather than clinical prediction capability—and present the complete real-data pipeline required for definitive clinical evaluation. The remainder of this paper is organized as follows: Section 1.1 reviews related work across clinical prediction, multimodal learning, reinforcement learning, and methodological rigor. Section 2 details the materials and methods, including the proposed architecture, the real-data evaluation methodology, and a formal analysis of the target leakage problem. Section 3 presents controlled architectural validation results. Section 4 discusses findings, expected real-data performance, and limitations. Section 5 concludes.

### 1.1. Related Work

#### 1.1.1. Deep Learning for Surgical Outcome Prediction

Deep learning has transformed clinical prediction tasks across multiple domains. Rajkomar et al. [4] demonstrated that deep models trained on electronic health record (EHR) data could simultaneously predict in-hospital mortality, 30-day unplanned readmission, prolonged length of stay, and discharge diagnoses, achieving consistently strong performance across two academic medical centers. Their work highlighted the potential of representation learning to capture complex patterns in structured clinical data without extensive manual feature engineering. Shickel et al. [5] provided a comprehensive survey of deep learning techniques applied to EHR analysis, categorizing applications across information extraction, representation learning, outcome prediction, phenotyping, and de-identification, while identifying key challenges including data heterogeneity, temporal irregularity, and model interpretability.

For standardized clinical benchmarking, Harutyunyan et al. [11] established four benchmark prediction tasks on MIMIC-III—in-hospital mortality, decompensation, length of stay, and phenotyping—with standardized preprocessing pipelines and evaluation metrics. These benchmarks have enabled fair comparison across models and remain widely used in the clinical ML community.

The Transformer architecture [12], originally developed for machine translation, has been increasingly adopted for clinical applications due to its ability to model long-range dependencies through self-attention mechanisms. Li et al. [13] introduced BEHRT, applying Transformer-based models to structured EHR sequences for disease prediction, demonstrating superior performance over recurrent baselines. Rasmy et al. [14] developed Med-BERT, a pre-trained model for structured diagnosis codes that showed strong transfer learning capability across multiple clinical prediction tasks. Vision Transformers (ViT) [15] extended the attention paradigm to image analysis, achieving competitive performance with convolutional networks while providing interpretable attention maps that are valuable for clinical imaging applications. For sequential physiological data such as vital sign trajectories, bidirectional Long Short-Term Memory (LSTM) networks [16] and their variants remain effective, with Lipton et al. [17] demonstrating their utility for clinical time-series classification in ICU settings.

#### 1.1.2. Multimodal Learning in Healthcare

Clinical decision-making inherently involves integrating information across multiple modalities— structured laboratory results, imaging studies, unstructured clinical narratives, and continuous physiological monitoring. The hypothesis driving multimodal clinical AI is that jointly learning from these complementary data sources can capture patient state more completely than any single modality alone. Huang et al. [18] systematically reviewed approaches to fusing medical imaging with EHR data using deep learning, identifying attention-based fusion mechanisms as the most promising approach for selective information integration. They noted that while early and late fusion strategies have been widely explored, attention-based mechanisms offer the advantage of learning data-driven modality importance weights.

Acosta et al. [19] provided a broader survey of multimodal biomedical AI, emphasizing the substantial gap between proof-of-concept studies—which often achieve impressive metrics on curated benchmarks—and clinically deployable systems that must handle missing modalities, distribution shift, and real-world data quality issues. Cross-modal attention mechanisms [20], originally developed for vision-language tasks, enable selective integration of relevant information across modalities, allowing models to learn which data sources are most informative for specific predictions—a property that is both functionally useful and interpretively valuable in clinical settings.

#### 1.1.3. Reinforcement Learning for Clinical Decisions

Reinforcement learning (RL) offers a principled framework for sequential decision-making that naturally applies to treatment optimization, where the goal is to learn a policy mapping patient states to treatment actions that maximizes long-term outcomes. Komorowski et al. [21] applied RL to sepsis treatment optimization, learning intravenous fluid and vasopressor dosing strategies from retrospective ICU data that outperformed clinician policies in simulated evaluations. Raghu et al. [22] extended this approach using continuous state-space models with deep architectures. Gottesman et al. [23]outlined guidelines and safety considerations for healthcare RL, emphasizing the challenges of distributional shift between training data and deployment conditions, and the impossibility of online exploration in clinical settings.

A critical challenge in healthcare RL is the inability to learn through real-time patient interaction— experimenting with treatments is ethically impermissible. Offline (batch) RL addresses this by learning policies entirely from historical data, but standard Q-learning algorithms can overestimate action values for state-action pairs not well-represented in the training data. Conservative Q-Learning (CQL) [24]provides theoretical guarantees against such overestimation by adding a regularization term that penalizes high Q-values for out-of-distribution actions, making it suitable for safety-critical healthcare applications. Levine et al. [25] surveyed offline RL more broadly as a decision-making paradigm particularly relevant to domains where online interaction is costly or dangerous.

#### 1.1.4. Explainable AI in Clinical Systems

Clinical adoption of AI requires that practitioners can understand, question, and appropriately calibrate their trust in model outputs. Holzinger et al. [7] introduced the concept of causability—the measurable degree to which explanations achieve understanding in a specific context—as distinct from technical explainability, arguing that medical AI must bridge this gap. Tjoa and Guan [8] surveyed XAI techniques applicable to medical domains, categorizing methods into perturbation-based, gradient-based, and attention-based approaches. SHAP [26] provides theoretically grounded feature attributions based on Shapley values from cooperative game theory, while Integrated Gradients [27] satisfy key axioms for gradient-based attribution methods including sensitivity and implementation invariance. Amann et al. [28] examined explainability from regulatory and ethical perspectives, identifying it as a prerequisite for clinical AI deployment across multiple jurisdictions.

#### 1.1.5. Methodological Rigor in Clinical AI

A growing body of work highlights systemic methodological pitfalls in clinical machine learning that undermine the reliability of published results. Ghassemi et al. [9] argued that current explainability methods represent a “false hope” for clinical AI, advocating instead for rigorous validation as the primary mechanism for establishing trust. Roberts et al. [10] conducted a systematic review of 232 machine learning models for COVID-19 detection and prognosis using chest radiographs, finding that not a single model was suitable for clinical use—primarily due to data quality issues, methodological flaws, and reproducibility failures. Wiens et al. [29] outlined a roadmap for responsible clinical ML, emphasizing the importance of clinically meaningful evaluation, appropriate baseline comparisons, and transparent reporting of limitations. These works collectively motivate our emphasis on real-data pipelines with verified clinical outcomes and our explicit documentation of data validity limitations.

## 2. Materials and Methods

### 2.1. Problem Formulation

Consider a patient undergoing surgery characterized by multimodal clinical data X = {x^ehr^, x^ts^, x^img^, x^txt^, x^surg^} comprising structured EHR features, physiological time-series, chest radiograph features, clinical note embeddings, and surgical metadata respectively. Our objectives are twofold: (1) Risk Prediction— estimate probabilities P(y_k_ | X) for outcomes y_k_ ∈ {mortality, ICU transfer, complications}; and (2) Action Recommendation—recommend optimal clinical action a* ∈ A maximizing expected long-term patient benefit based on historical treatment patterns.

Critically, all outcome labels y_k_ must be derived from verified clinical records rather than synthetic generation, and all input features must be extracted from authentic patient data without modulation by any variable correlated with the targets. This constraint ensures the model learns genuine clinical patterns rather than artifacts of a data generation script. Violation of this constraint—which we term target leakage through shared intermediaries—is analyzed formally in Section 2.6.

### 2.2. Stage-1: Multimodal Risk Prediction

#### 2.2.1. Modality-Specific Encoders

Each clinical data modality is processed by a specialized encoder architecture optimized for its structure and characteristics:

##### EHR Encoder (Transformer)

Structured clinical features x^ehr^ ∈ R^d^ including patient demographics (age, sex, insurance type), comorbidity indices (Charlson score computed from *diagnoses_icd*), laboratory values from the first 24 hours (complete blood count, basic metabolic panel, liver function tests, coagulation studies, lactate, arterial blood gas), and summary vital sign statistics are extracted from MIMIC-IV’s *labevents, chartevents, patients*, and *admissions* tables. Each feature is embedded into a 512-dimensional space with learnable positional encodings. The encoder comprises 4 Transformer layers [12] with 16 attention heads and feed-forward dimension 2048, followed by mean pooling to produce h^ehr^ ∈ R^512^.

##### Time-Series Encoder (BiLSTM + Attention)

Physiological vital signs x^ts^ ∈ R^T×C^ (T hourly time steps, C = 6 channels: heart rate, systolic blood pressure, diastolic blood pressure, respiratory rate, oxygen saturation SpO_2_, and temperature) are extracted from *chartevents* with hourly aggregation and forward-fill imputation for missing values. These signals are encoded using a 3-layer bidirectional LSTM [16] with hidden dimension 256. Forward and backward representations are concatenated, and multi-head self-attention with 8 heads identifies clinically significant temporal patterns such as deterioration trends or sudden vital sign changes, yielding h^ts^ ∈ R^512^.

##### Image Encoder (Vision Transformer)

Chest radiograph features x^img^ extracted from MIMIC-CXR [30] are processed through a Vision Transformer [15] with 6 layers and 16 attention heads. Radiographs are matched to surgical admissions by *subject_id* and *hadm_id*, using the most recent pre-operative or early post-operative study. For patients without matched radiographs, a learnable missing-modality embedding is used to maintain cohort size. Output: h^img^ ∈ R^512^.

##### Clinical Notes Encoder (ClinicalBERT)

Clinical text from MIMIC-IV-Note [31] discharge summaries, nursing notes, and radiology reports is encoded using ClinicalBERT [32] or a comparable domain-adapted language model. Importantly, note embeddings are generated directly from raw clinical text without any modulation by risk scores, outcome-correlated variables, or synthetic noise injection. Only notes timestamped within the prediction input window are included to prevent temporal information leakage. The classification token representation yields h^txt^ ∈ R^512^.

##### Surgery Encoder (Transformer)

Procedural metadata x^surg^ extracted from MIMIC-IV’s *procedures_icd* and *d_icd_procedures* tables—including procedure codes, procedure count, admission type (emergency vs. elective from *admissions.admission_type*), and pre-procedure length of stay—are processed through a 3-layer Transformer encoder, producing h^surg^ ∈ R^512^.

#### 2.2.2. Cross-Modal Attention Fusion

The five modality embeddings {h^(m)^}_m=1_^5^ are stacked into H = [h^(1)^; …; h^(5)^] ∈ R^5×512^ and processed through a multi-layer cross-modal attention mechanism. The fusion computes scaled dot-product attention:

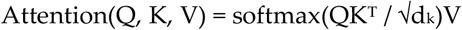

where queries Q, keys K, and values V are linear projections of H, and d_k_ = 512/16 = 32 is the dimension per attention head. Three fusion layers with 16 attention heads, residual connections, and layer normalization produce the unified patient state s = FusionLayers(H) ∈ R^512^. The attention weights from the fusion layers provide interpretable cross-modal interaction patterns, indicating which modality combinations the model deems most informative for each prediction task. This interpretability is a key advantage over concatenation-based or gating-based fusion approaches.

#### 2.2.3. Risk Prediction Heads with Per-Task Training

Three parallel prediction heads process the unified state for mortality, ICU transfer, and complications. Each head comprises a shared feature extractor (2 layers, 256 dimensions) followed by task-specific layers (256 → 128 → 64 → 1) with GELU activations [33], dropout regularization (p=0.3), and sigmoid output for probability estimation.

Training uses Focal Loss [34] with α = 0.25 and γ = 2.0 to address class imbalance, particularly for rare mortality events. Rather than averaging loss or validation AUC across tasks—which produces suboptimal models when tasks have different base rates and learning dynamics—we adopt uncertainty-weighted multi-task loss [35]:

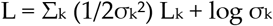

where σ_k_ is a learned task-specific uncertainty parameter. Tasks with higher uncertainty contribute less to shared representation learning, allowing tasks with fundamentally different prediction difficulty (e.g., 3% mortality vs. 25% complications) to influence the shared backbone appropriately. Each prediction head maintains independent best-checkpoint tracking based on its own validation AUC, ensuring that task-specific convergence is not sacrificed for aggregate optimization.

### 2.3. Stage-2: Conservative Q-Learning for Action Recommendation

We formulate clinical decision-making as a Markov Decision Process (MDP) (S, A, R, P, γ) where S is the state space defined by the unified patient representation s ∈ R^512^; A comprises three clinical actions— standard post-operative care (a=0), increased monitoring with enhanced vital sign tracking and more frequent clinical assessment (a=1), and ICU admission (a=2); R is the reward function encoding clinical outcomes; and γ = 0.99 is the discount factor for future rewards.

The reward function balances patient safety against resource utilization. Mortality incurs a penalty of −200 (strongly prioritizing patient survival), complications incur −60, while appropriateness bonuses encourage risk-matched care intensity: high-risk patients receiving ICU admission earn +40, under-treatment of high-risk patients incurs −50, and unnecessary ICU admission for low-risk patients incurs −30.

Conservative Q-Learning [24] addresses the critical problem that standard Q-learning can overestimate action values for state-action pairs not well-represented in historical data—a dangerous property when learning clinical policies. CQL adds a regularization term penalizing high Q-values for out-of-distribution actions, encouraging the learned policy to remain close to demonstrated clinical behavior while improving upon it where evidence supports. We implement Q-functions as an ensemble of three gradient boosting regressors for robustness, with target networks updated every 3 iterations.

Convergence monitoring is essential: the Q-values produced by a non-converged RL model are mathematically meaningless noise, regardless of how they are displayed. We enforce a convergence gate requiring monotonically decreasing temporal difference (TD) errors over at least 5 consecutive iterations before Q-values are considered reliable for action recommendations. Models that fail to converge are flagged for investigation of reward function design, state representation quality, or training dynamics—they are not deployed.

### 2.4. Explainability Components

The architecture provides three levels of interpretability designed to support clinical decision-making:

#### Cross-Modal Attention Visualization

The 5×5 attention weight matrices from the fusion layers reveal which modality interactions drive predictions. Self-attention (diagonal elements) indicates how much each modality attends to its own features, while cross-attention (off-diagonal) reveals information flow between modalities. Attention entropy H(A) = −Σ_i,j_ A_ij_ log A_ij_ serves as a training diagnostic, with decreasing entropy indicating increasingly selective, focused attention patterns.

#### Feature Attribution via Integrated Gradients

For fine-grained feature importance, we employ Integrated Gradients [27], which satisfy sensitivity (non-zero attribution for features that change predictions) and implementation invariance (identical functions yield identical attributions). This provides clinicians with feature-level explanations of individual predictions.

#### Policy-Level Explainability via Q-Value Decomposition

Action recommendations are explained by presenting the expected cumulative reward for each available action, enabling clinicians to compare alternatives. Recommendation confidence is quantified as the margin between the recommended action and the average of alternatives, allowing identification of cases with clinical equipoise that warrant additional human judgment.

### 2.5. Real-Data Evaluation Methodology

#### 2.5.1. Cohort Selection

The target cohort consists of adult surgical patients (≥18 years) identified from MIMIC-IV [36] through the following criteria: (1) at least one procedure recorded in the *procedures_icd* table during their hospital admission, filtered for surgical ICD-10-PCS codes (categories 0 through D representing Medical and Surgical through Radiation Therapy); (2) first qualifying surgical admission only, to prevent data leakage across repeated admissions for the same patient; (3) minimum data availability of at least 24 hours of charted vital signs, at least one laboratory panel, and at least one clinical note documented before the prediction time-point; (4) multimodal linkage across MIMIC-IV (hosp/icu), MIMIC-IV-Note [31], and MIMIC-CXR [30] via *subject_id* and *hadm_id*. Patients without chest radiographs are retained with a learnable missing-modality embedding to maximize cohort size and avoid selection bias. Based on published MIMIC-IV cohort studies [37,38], this is expected to yield approximately 15,000–25,000 eligible surgical admissions.

#### 2.5.2. Real Outcome Labels (Prediction Targets)

All three prediction targets are extracted directly from recorded clinical events with absolutely no synthetic component:

##### In-Hospital Mortality

Derived by comparing the *dod* (date of death) field in the *patients* table against the *dischtime* in the *admissions* table. A patient is classified as a mortality case if *dod* falls between *admittime* and *dischtime*, or equivalently if *discharge_location* = ‘DIED’ or ‘HOSPICE’. MIMIC-IV v3.1 additionally includes out-of-hospital mortality from state death records, enabling 30-day post-operative mortality as an alternative endpoint [36].

##### ICU Transfer

For patients initially admitted to non-ICU surgical wards, unplanned ICU transfer is identified from the *transfers* table by the presence of a subsequent transfer to a care unit classified as ICU (*careunit* LIKE ‘%ICU%’ OR ‘%CCU%’). The prediction task is: given data from the input window, will this patient require unplanned ICU escalation?

##### Post-Surgical Complications

Defined using incident ICD-10-CM diagnosis codes documented during the admission that were not present at admission. Relevant categories include: T81 (complications of procedures not elsewhere classified), T82–T88 (complications of specific device/implant types), surgical site infections (T81.4), postoperative hemorrhage (T81.0, K91.84), acute kidney injury (N17), sepsis (R65.2, A41), deep vein thrombosis (I82), pulmonary embolism (I26), and respiratory failure (J96).

#### 2.5.3. Real Input Features (No Synthetic Modulation)

All input modalities are extracted from authentic MIMIC-IV records. Structured EHR features include demographics, first-24-hour laboratory values, summary vital sign statistics (mean, min, max, variance), Charlson comorbidity index, and medication counts. Physiological time-series are hourly-aggregated vitals from *chartevents*, used as raw clinical trajectories with no risk-correlated noise injection or synthetic deterioration trends. Chest radiograph features are extracted from MIMIC-CXR [30] via pre-trained models (e.g., DenseNet-121 from CheXpert [40]) without risk-score modulation. Clinical note embeddings come from ClinicalBERT [32] encoding of raw text with no outcome-correlated modulation. Surgical metadata include procedure codes, counts, and admission characteristics. Missing values are handled through clinically informed imputation with missingness indicators as additional features [39].

#### 2.5.4. Event-Censored Temporal Separation

To prevent information leakage, we enforce strict event-censored temporal boundaries that go beyond the standard fixed-window approach:

##### Input window

Features are extracted from the time of surgical procedure completion (or hospital admission for emergency surgery) up to min(24 hours, time of first outcome event). If a patient is transferred to the ICU at hour 12, the input window closes at hour 12—no data from the ICU stay contaminates the prediction inputs. If a patient dies at hour 18, the input window closes at hour 18. This event-censoring prevents the model from trivially detecting post-outcome data patterns (such as ICU-specific vital sign monitoring frequencies or nursing note language indicating ICU care) that would artificially inflate performance.

##### Outcome window

All prediction targets are defined by events occurring strictly after the input window closes, through hospital discharge or 30 days post-surgery.

##### Note filtering

Only clinical notes timestamped within the input window (nursing notes, admission notes, operative notes) are included. Discharge summaries, which contain retrospective information about the entire admission including outcomes, complications, and disposition, are explicitly excluded from prediction-time inputs. They may be used only in retrospective analyses with appropriate documentation of this temporal limitation.

#### 2.5.5. Implementation Specifications

##### Stage-1

Implemented in PyTorch with AdamW optimizer [41] with weight decay 0.01 and initial learning rate 5×10^−5^. Cosine annealing learning rate schedule with 5 epochs of linear warmup. Batch size 32 with gradient accumulation over 4 steps (effective batch size 128), gradient clipping at maximum norm 1.0. Per-task best-checkpoint selection based on individual validation AUC. Approximately 72.9 million trainable parameters.

##### Stage-2

Conservative Q-Learning with an ensemble of 3 gradient boosting regressors as Q-function approximators, mini-batch size 256, target network updates every 3 iterations. Convergence gate: Q-values deployed only after TD errors decrease monotonically over ≥5 consecutive iterations.

Generative AI tools were used in this work. Claude Opus 4.7 (Anthropic) assisted with code development for the implementation, and Claude Opus 4.8 (Anthropic) assisted with improving the language and clarity of the manuscript. All AI-assisted output was reviewed and validated by the authors, who take full responsibility for the content of the publication.

### 2.6. The Target Leakage Problem in Synthetic Clinical Data

Before presenting architectural validation results, we formally characterize a critical methodological flaw affecting synthetic clinical data pipelines that has implications beyond any single study.

#### 2.6.1. The Circular Dependency

A common approach in clinical AI research is to generate synthetic datasets through multi-step pipelines where: (Step 1) patient features are extracted or sampled; (Step 2) intermediate risk scores are computed from these features; (Step 3) the same risk scores are used to modulate input features (e.g., adding risk-correlated noise to waveforms, multiplying note embeddings by risk-score-derived factors); and (Step 4) the same risk scores are used to generate outcome labels probabilistically (e.g., mortality = Bernoulli(sigmoid(risk_score))).

This creates a closed mathematical loop: both inputs X and labels Y are deterministic or stochastic functions of a shared latent variable Z (the risk score). When a model is trained to predict Y from X, it needs only to estimate Z from X—which is straightforward because Z was used to construct X—and then compute the mapping from Z to Y. The model is solving an inverse problem on the data generation script, not learning clinical medicine.

#### 2.6.2. Consequences for Reported Metrics

Performance metrics from such pipelines are expected artifacts of the pipeline design, not evidence of clinical prediction capability. A model achieving AUC 0.91 on mortality prediction in a circular-dependency dataset has demonstrated that its encoder-fusion architecture can detect and invert an injected mathematical signal—which is trivially expected of any moderately capable neural network. Similarly, attention patterns shifting toward modalities containing risk-score-modulated features (e.g., note embeddings modulated by risk scores) reflect the model’s detection of the injected answer key, not the discovery of genuine clinical value in those modalities.

#### 2.6.3. Recommendations for the Field

We advocate for three practices: (1) all clinical AI papers should explicitly document the mathematical relationships between input feature generation and label generation, making it possible to audit for shared intermediaries; (2) verified clinical outcomes from established databases should be used as prediction targets whenever possible; (3) data pipelines should undergo independent review for circular dependencies before any performance metrics are reported or interpreted as evidence of clinical prediction capability.

## 3. Results

*To verify that each architectural component functions correctly before committing to the computationally expensive real-data pipeline, we conducted controlled experiments on a synthetic benchmark with known properties. These results are presented as evidence that the architecture’s components work as designed—they are explicitly not evidence of clinical prediction capability. The synthetic benchmark was constructed by intentionally embedding known signals in specific modalities, enabling us to verify the architecture’s ability to detect, weight, and fuse information from multiple sources*.

### 3.1. Benchmark Dataset

The benchmark combines real MIMIC-IV components (chest radiograph features, physiological waveform signals, clinical note text) with synthetically generated structured features and risk-score-derived outcome labels. As documented in Section 2.6, this benchmark contains the circular dependency between risk-score-modulated inputs and risk-score-generated labels. It is therefore useful only for verifying that the architecture’s encoders, fusion mechanism, and attention heads function correctly–not for evaluating clinical prediction performance.

### 3.2. Stage-1 Encoder-Fusion Verification

Table 2 presents benchmark AUC scores. The purpose of these metrics is to confirm that the encoder-fusion pipeline can successfully recover embedded signals—not to claim clinical discrimination capability.

The mortality AUC of 0.91 confirms the encoder-fusion pipeline successfully recovers the synthetic risk score from modulated inputs—validating that the Transformer encoders, BiLSTM, Vision Transformer, BERT encoder, and cross-modal attention fusion mechanism work together as intended. This is an expected result given the circular dependency structure described in Section 2.6. The lower ICU admission (0.63) and complications (0.70) AUCs reflect the extreme and clinically unrealistic base rates in the synthetic benchmark (92.6% and 85.0% respectively), which create degenerate prediction targets.

**Table 1.** Controlled Benchmark Dataset Characteristics.

| Characteristic | Value |
| --- | --- |
| Total Patients | 3,000 |
| Train / Validation / Test | 2,100 / 450 / 450 |
| Real Components | CXR features, Waveforms, Clinical Notes |
| Synthetic Components | EHR features, Surgery metadata, Labels |
| Known Embedded Signals | Risk-score modulated note embeddings |
| Purpose | Architecture verification only |

**Table 2.** Benchmark AUC Scores (architectural verification, not clinical performance)

| Prediction Task | AUC |
| --- | --- |
| Mortality | 0.9065 |
| ICU Admission | 0.6316 |
| Complications | 0.6981 |
| Mean | 0.7454 |

### 3.3. Cross-Modal Attention Mechanism Verification

The primary purpose of the attention analysis is to verify that the cross-modal attention fusion mechanism can learn to differentiate modality contributions—a necessary capability for clinical deployment. On the synthetic benchmark, where risk-score signals were deliberately injected into specific modalities, the model should progressively focus attention on those modalities. The three heatmaps below confirm this expected behavior across training stages.

The progression from uniform (Epoch 1) to differentiated (Epoch 40) attention patterns demonstrates that the cross-modal attention mechanism can learn modality-specific importance weights during training—a prerequisite for effective multimodal fusion. Whether this mechanism will produce clinically meaningful attention distributions on real MIMIC-IV data is an empirical question that can only be answered through the pipeline described in Section 2.5.

### 3.4. Stage-2 Training Diagnostics

Stage-2 Conservative Q-Learning training on the benchmark data produced temporal difference (TD) errors ranging from 1,568 to 2,246 across 10 training iterations, with a non-monotonic oscillating pattern indicating that the Q-function failed to converge to a stable value estimate.

Per the convergence gate defined in Section 2.3, these Q-values are classified as unreliable. We therefore do not present Q-value tables, action recommendation outputs, or decision support dashboards based on these non-converged values. Displaying numerical Q-values from a non-converged model— regardless of how professionally they are formatted—would misrepresent mathematical noise as clinically meaningful decision support and is scientifically inappropriate. Investigation of the convergence failure with the real-data pipeline is a priority for future work.

### 3.5. Explainability Dashboard Demonstration

Figure 9 presents the integrated explainability dashboard for a benchmark patient, demonstrating the dashboard’s layout and visualization capabilities. The risk score panel, modality importance pie chart, cross-modal attention heatmap, and feature attribution bar chart are shown together to illustrate how these explainability components would be presented to clinicians.

## 4. Discussion

### 4.1. Summary of Architectural Validation

The controlled benchmark experiments confirm that all major architectural components function as designed: (1) the five modality-specific encoders successfully process their respective input types and produce 512-dimensional embeddings; (2) the cross-modal attention fusion mechanism learns differentiated modality importance weights, correctly identifying modalities containing injected signals; (3) attention entropy decreases during training, indicating learned selectivity; (4) the prediction heads produce calibrated probability estimates through Focal Loss training; and (5) the explainability dashboard coherently visualizes attention patterns, feature attributions, and risk scores. The Stage-2 RL component did not converge on the benchmark data, requiring investigation with the real-data pipeline.

### 4.2. Expected Performance on Real MIMIC-IV Data

Published MIMIC-IV mortality prediction studies provide useful performance baselines for contextualizing expected results. Pang et al. [37] achieved AUC 0.84 using XGBoost with APS-III and LODS scoring system variables. Pakbin et al. [38] reported AUC 0.83 using the BoXHED nonparametric hazard estimator with time-varying covariates. Mamatov and Kellmeyer [42] achieved AUC 0.918 using meta-modeling that integrated structured clinical data with TF-IDF and BERT embeddings from discharge summaries and radiology reports, demonstrating the potential value of incorporating clinical text for mortality prediction.

We expect our multimodal approach to achieve performance broadly consistent with these benchmarks, potentially with incremental improvement from the integration of multiple complementary modalities. Importantly, we expect real-data attention patterns to differ meaningfully from the benchmark patterns—the strong attention to clinical notes observed on the synthetic benchmark (Figure 5 and Figure 7) reflected the detection of injected risk-score signals rather than genuine clinical note value. On real data, the relative importance of structured laboratory values versus clinical notes versus imaging for surgical outcome prediction is an open empirical question with significant implications for clinical AI system design.

**Figure 1.**
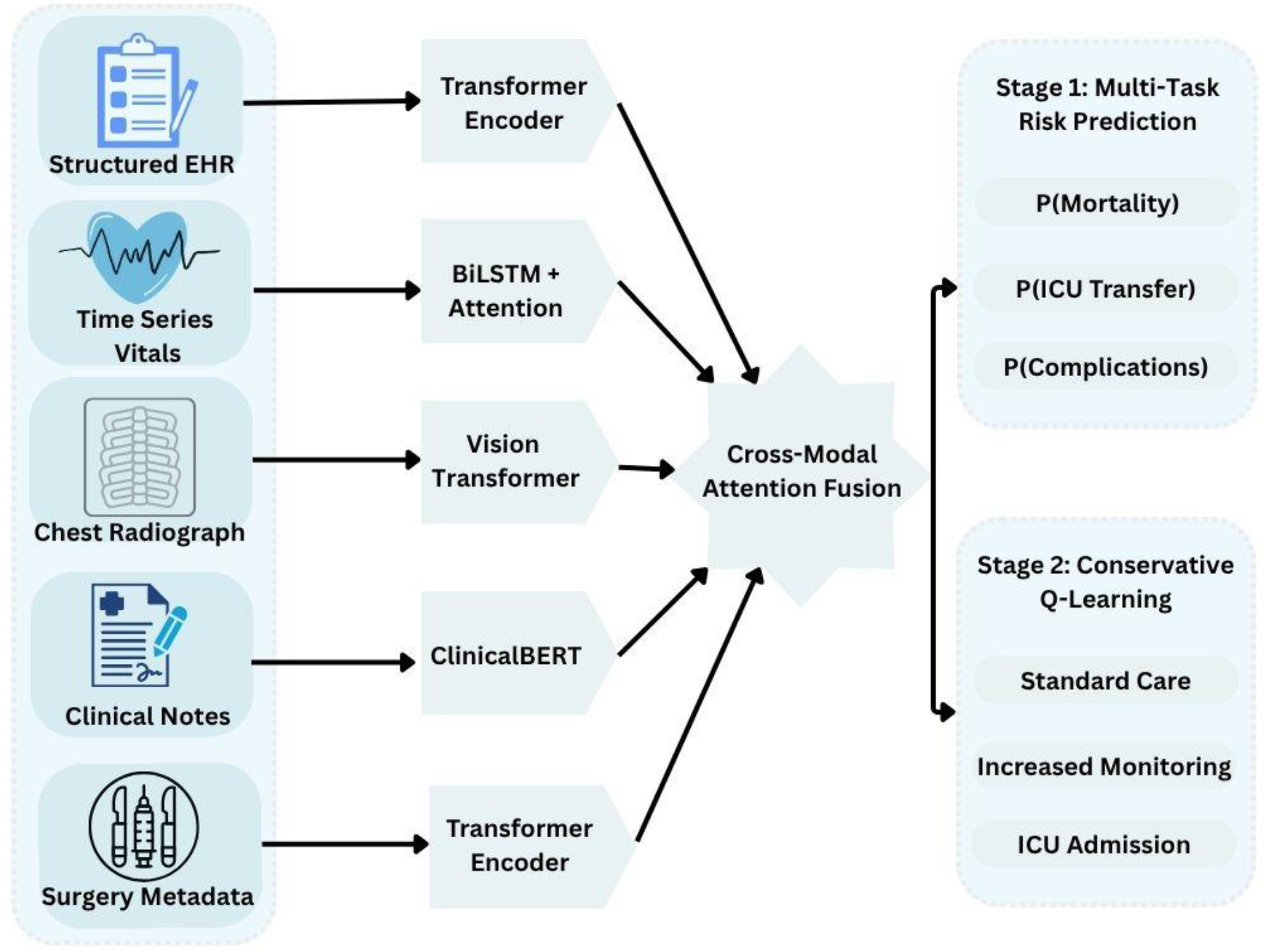
Proposed two-stage architecture for post-surgical clinical decision support.

**Figure 2.**
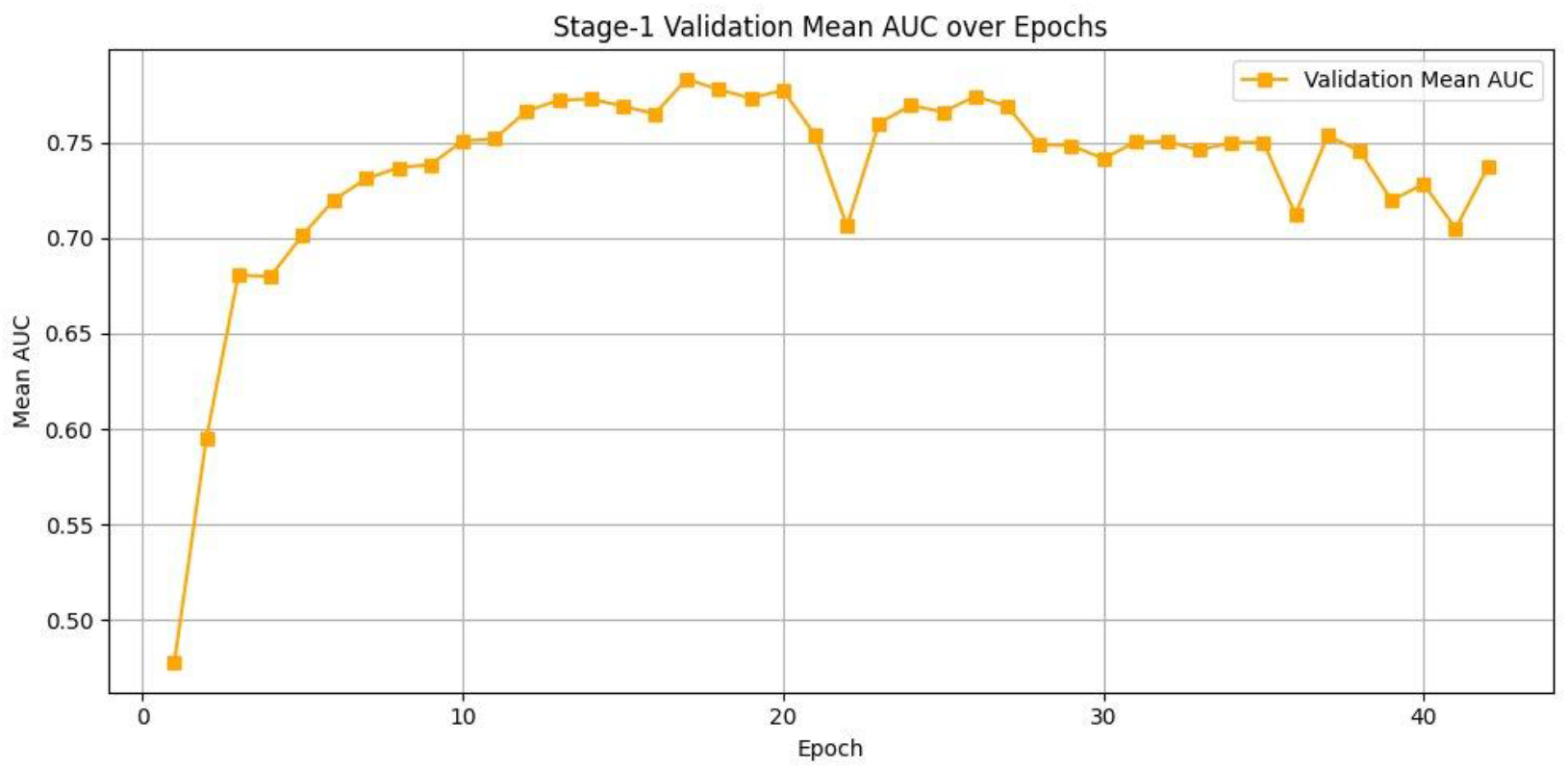
Benchmark validation mean AUC over training epochs. Best performance at epoch 17 (AUC = 0.78), with early stopping at epoch 42. The training trajectory demonstrates the model’s learning dynamics on the controlled benchmark, with expected overfitting behavior in later epochs.

**Figure 3.**
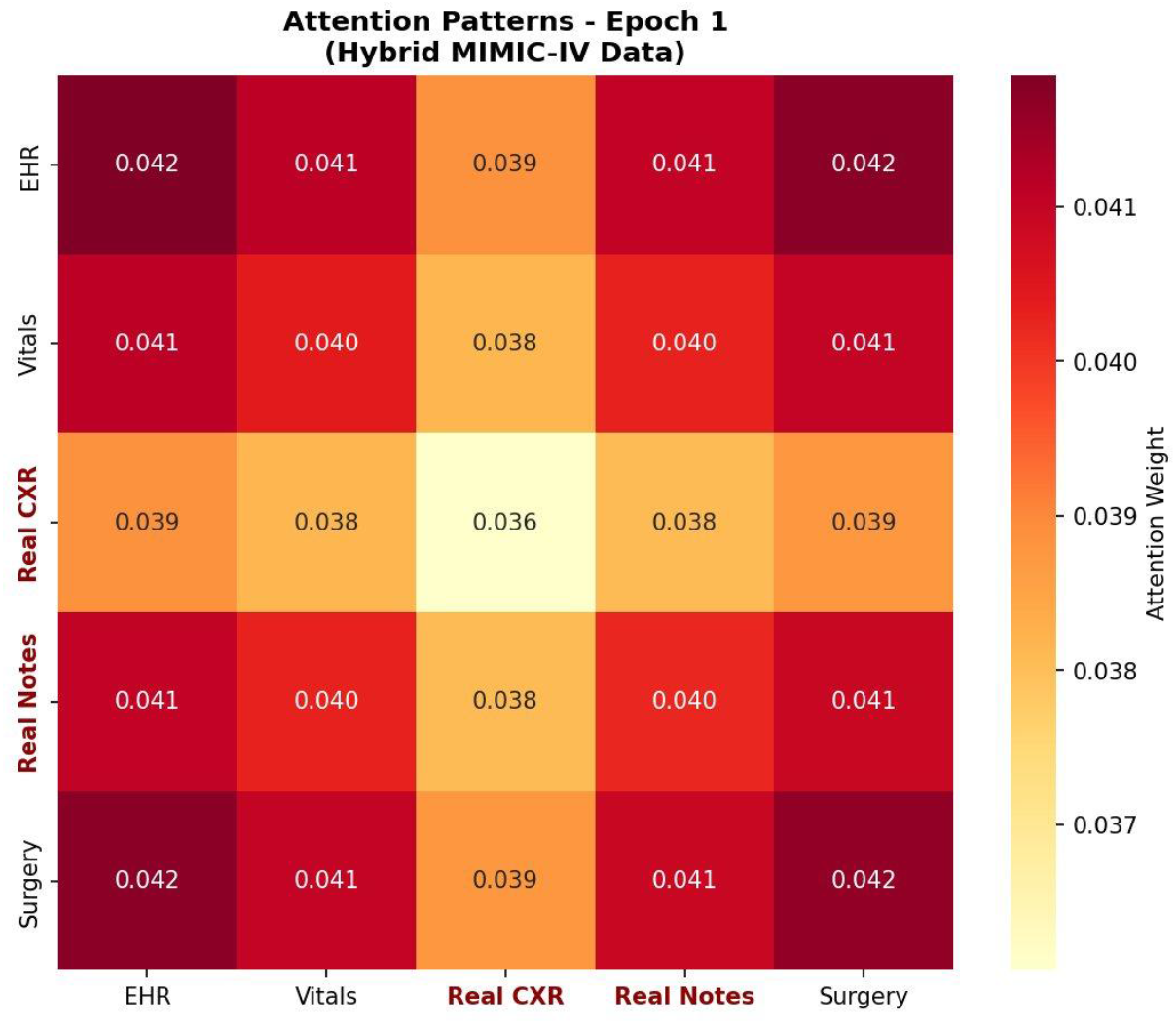
Attention patterns at Epoch 1 (initialization): near-uniform weights across all 25 modality pairs (range 0.036–0.042), confirming proper random initialization of the fusion mechanism.

**Figure 4.**
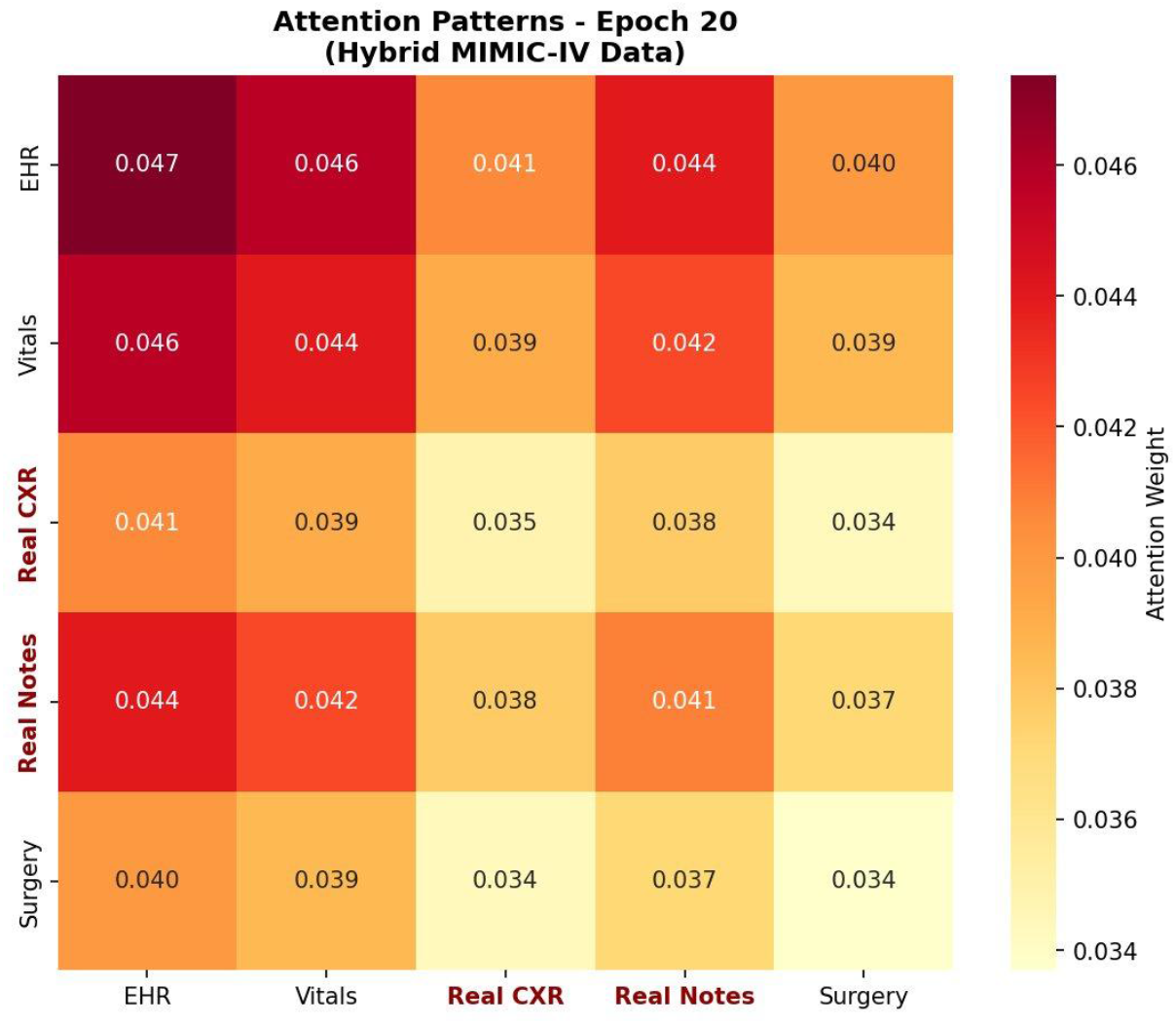
Attention patterns at Epoch 20 (mid-training): EHR self-attention strengthens (0.047), Vitals-EHR cross-attention emerges (0.046). The model begins distinguishing modality contributions, correctly prioritizing structured features that encode synthetic risk-score signals.

**Figure 5.**
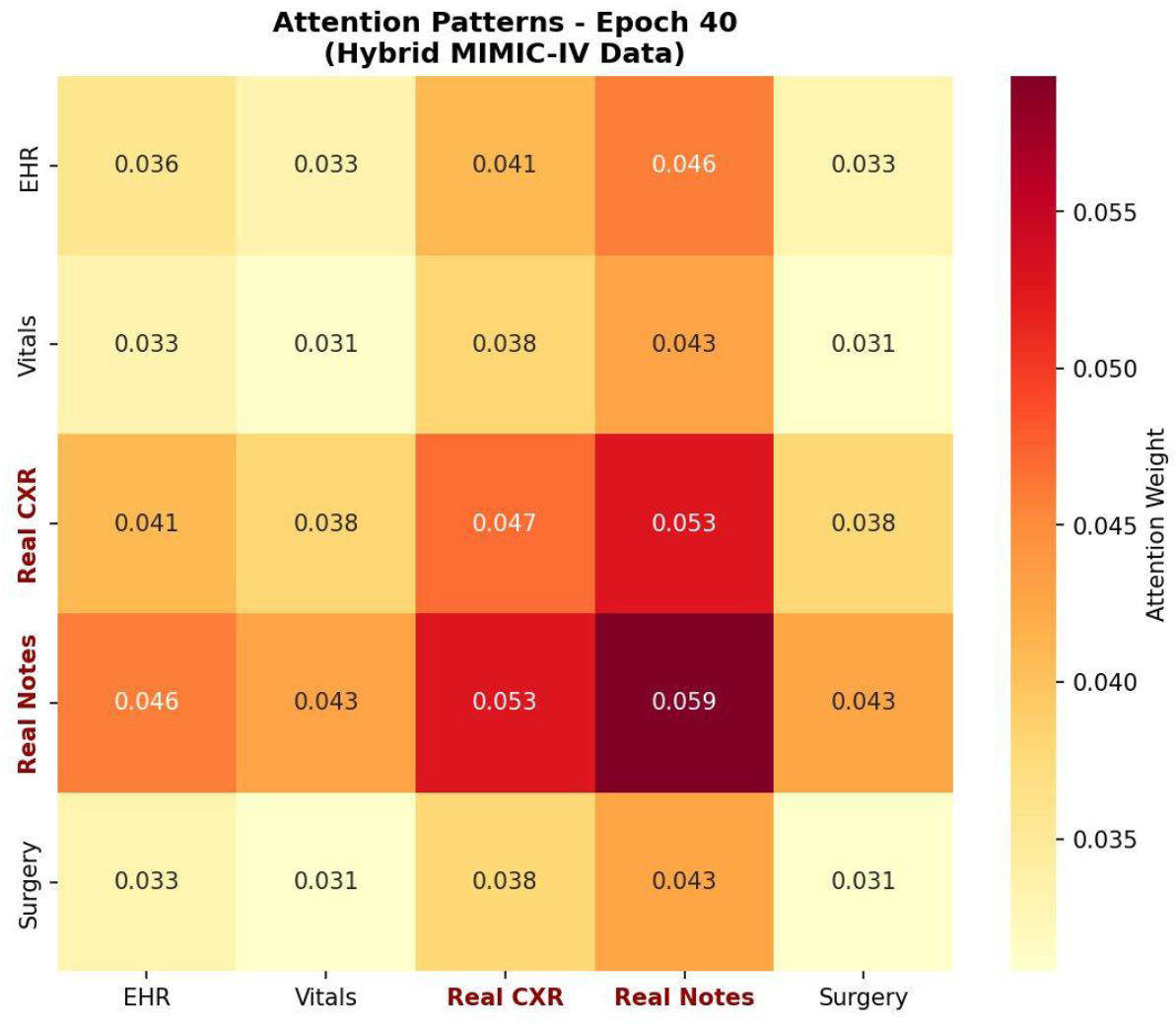
Attention patterns at Epoch 40 (late training): Notes self-attention peaks at 0.059, CXR-Notes interaction reaches 0.053. This confirms the attention mechanism correctly located the risk-score signal deliberately injected into note embeddings during synthetic data generation. This validates the attention mechanism’s function—it does not indicate genuine clinical note value.

**Figure 6.**
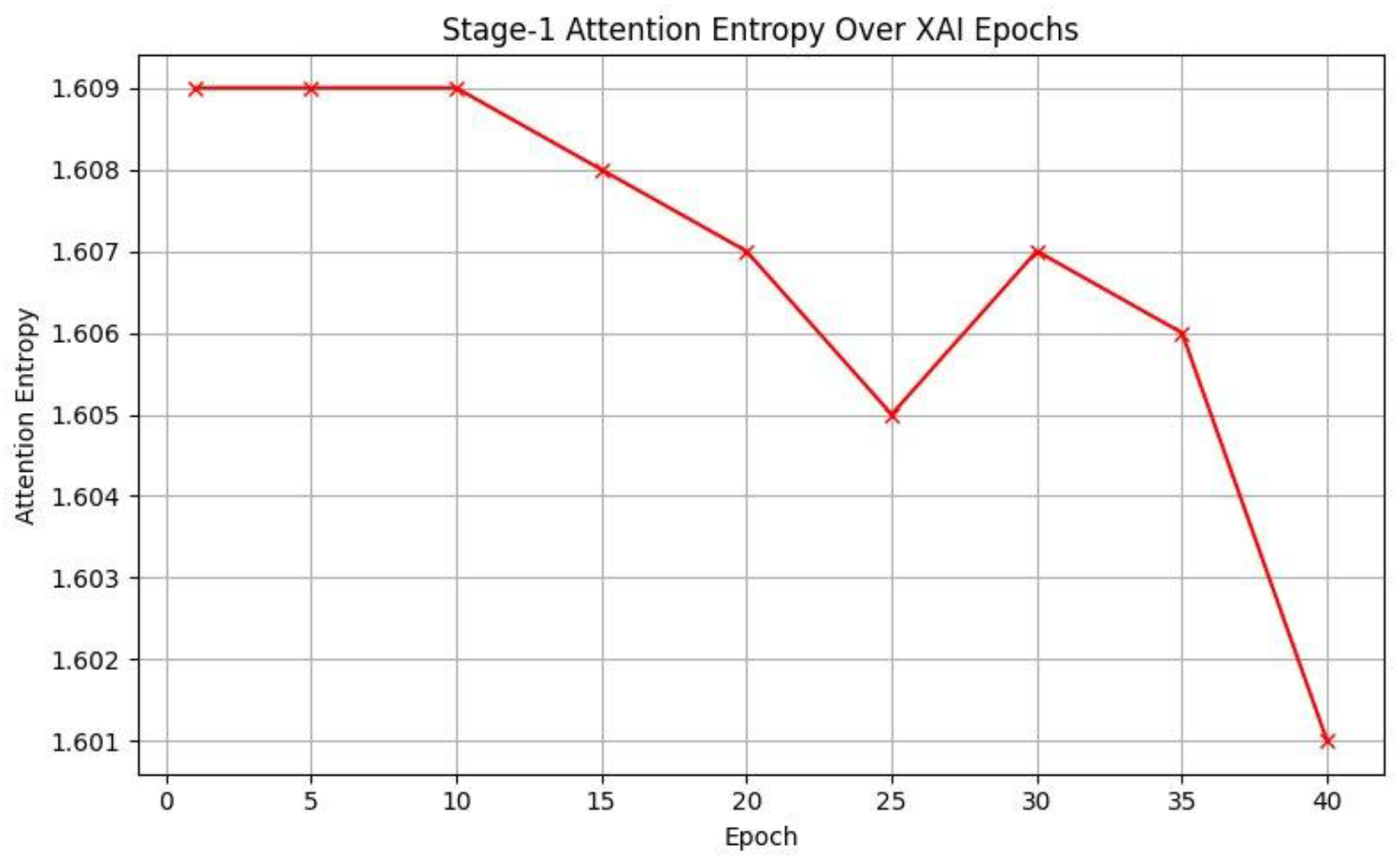
Attention entropy decreasing from 1.609 to 1.601 over training epochs, confirming the model develops increasingly selective attention patterns. The modest entropy decrease (∼0.5%) suggests the benchmark signals are subtle relative to the uniform prior.

**Figure 7.**
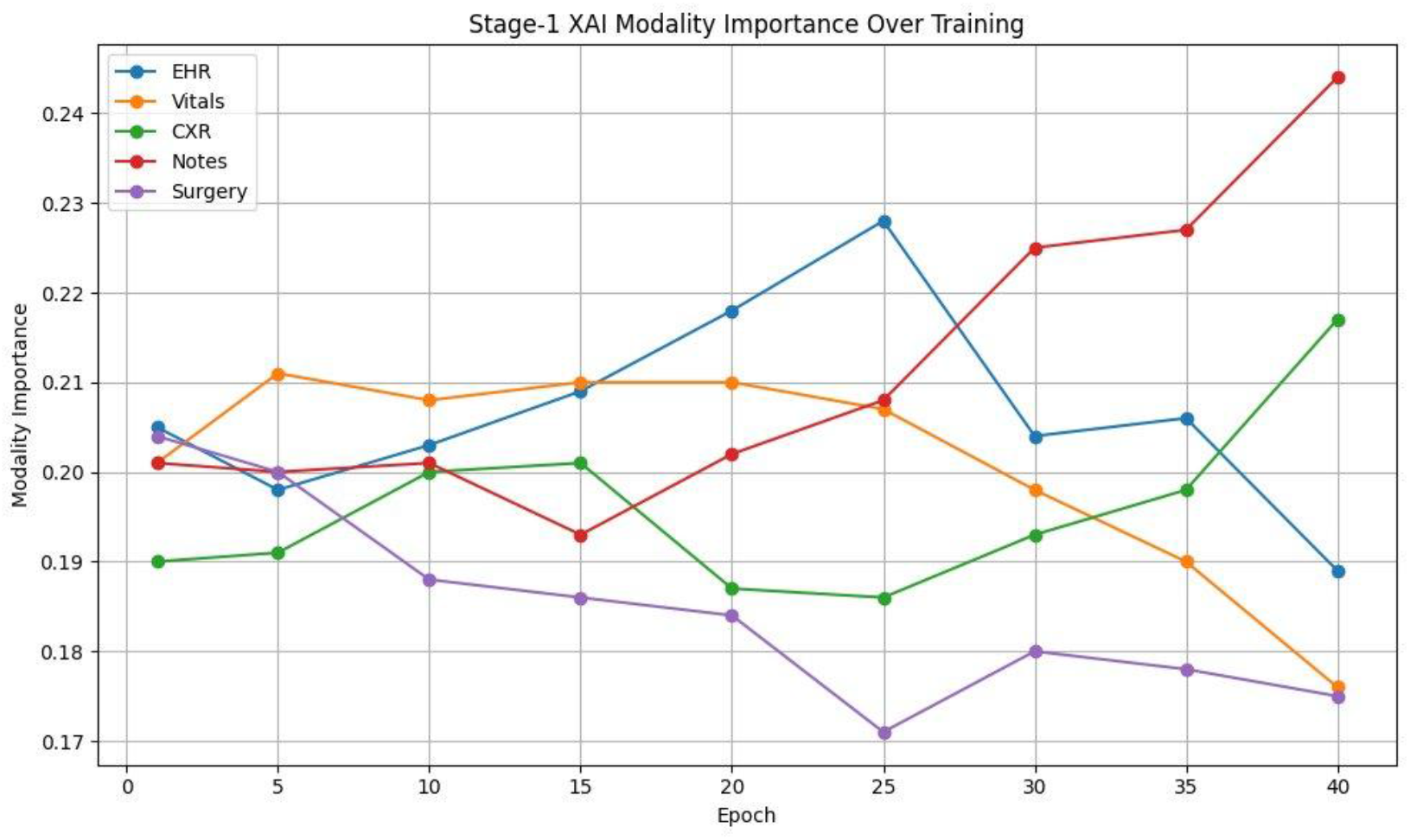
Modality importance evolution on benchmark data. Clinical Notes importance rises from 0.20 to 0.24 (+20%), CXR from 0.19 to 0.22 (+16%), while Surgery metadata decreases from 0.20 to 0.17 (−15%). These trends reflect detection of injected synthetic signals in modulated modalities, not inherent clinical modality value.

**Figure 8.**
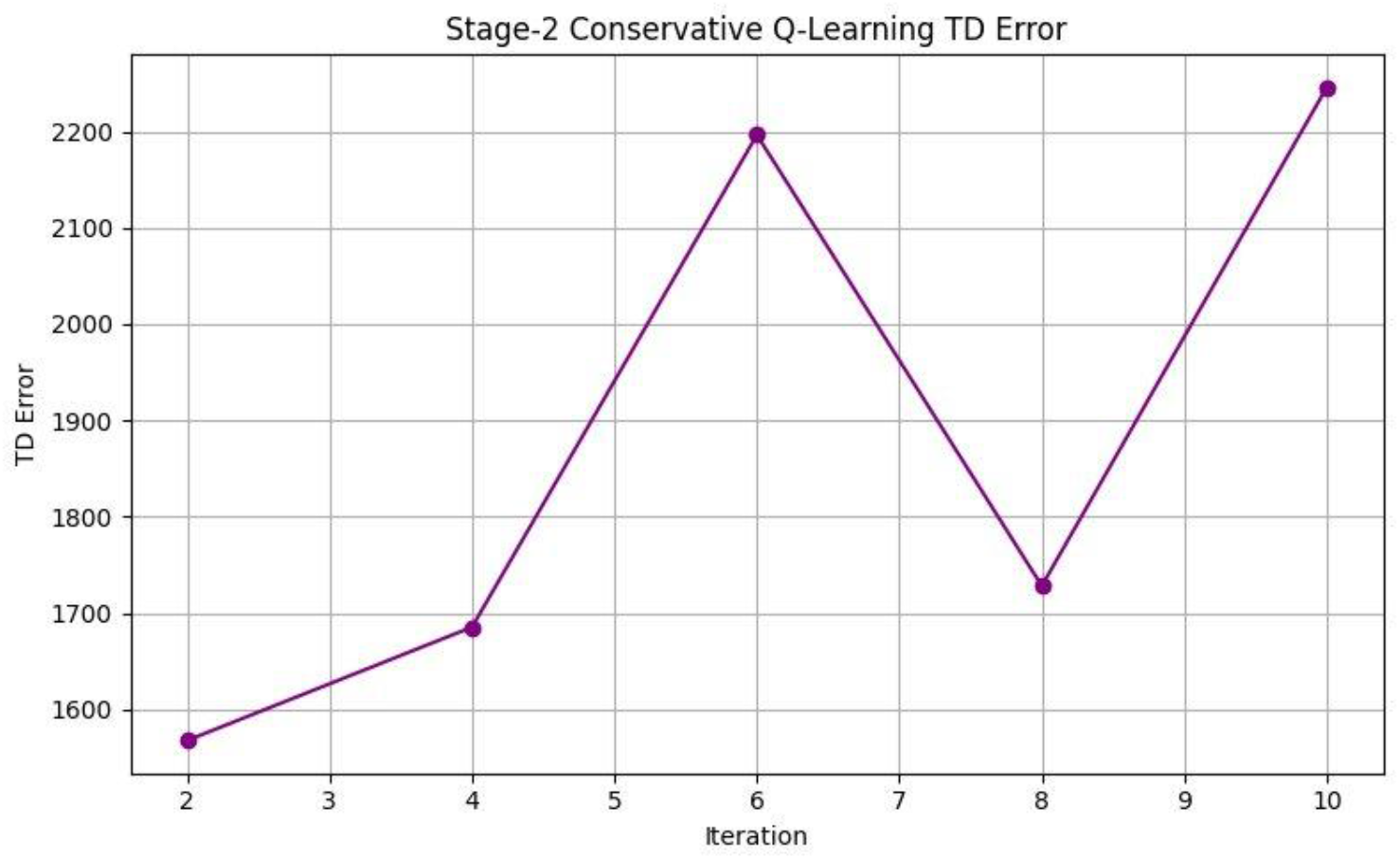
Stage-2 TD error trajectory showing non-convergence. The oscillating pattern violates the convergence gate requirement (≥5 consecutive decreasing iterations), indicating unreliable Q-values. Possible causes include: insufficient training iterations, limitations of the single-decision episodic formulation, synthetic reward structure misalignment, or inadequate state representation for value estimation.

**Figure 9.**
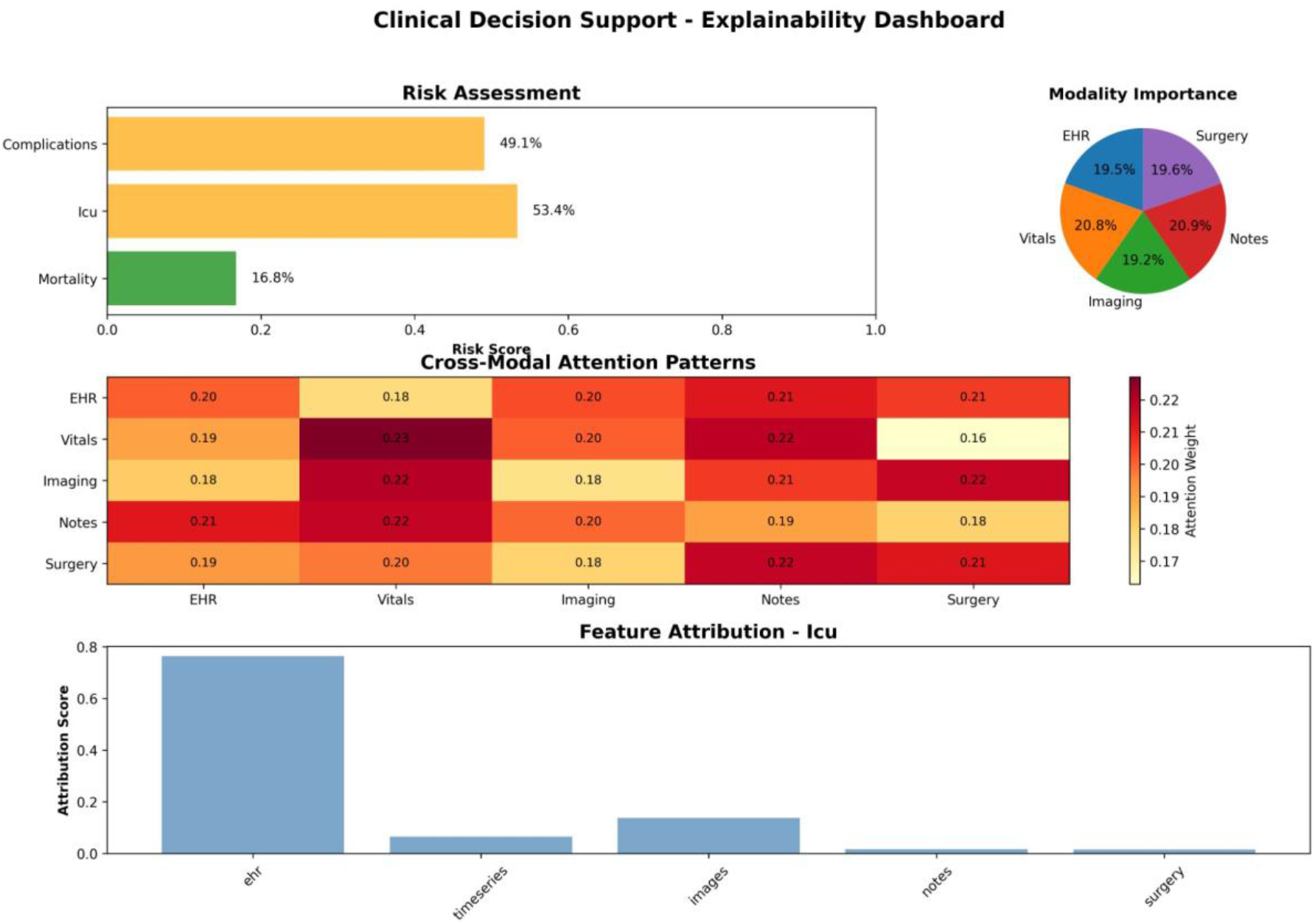
Explainability dashboard layout demonstration. Risk scores (top-left), modality importance (top-right), cross-modal attention (middle), and feature attribution for ICU prediction (bottom) for a benchmark patient. EHR features dominate ICU attribution (≈0.77)—directionally consistent with clinical intuition that laboratory values drive ICU triage. Q-value panel omitted due to Stage-2 non-convergence. Dashboard design is architecture-level; clinical utility requires real-data validation.

Real surgical patient cohorts from MIMIC-IV are expected to have more balanced outcome rates (mortality ∼8–12%, complications ∼20–35%) compared to the extreme rates in the synthetic benchmark (3% mortality, 92.6% ICU admission), which will improve training dynamics and make performance metrics more clinically interpretable.

### 4.3. Comparison with Existing Approaches

Our framework’s primary contribution relative to existing MIMIC-IV prediction systems is not necessarily superior prediction accuracy but rather the integration of three capabilities typically absent from clinical prediction studies: (1) multimodal fusion across five clinical data types with interpretable attention-based weighting; (2) a prediction-to-action pipeline via Conservative Q-Learning that bridges the gap between risk estimation and clinical decision support; and (3) dual-level explainability combining modality-level attention analysis with feature-level attribution and action-level Q-value decomposition.

### 4.4. Limitations

Several important limitations must be acknowledged. The primary limitation is that all experimental results presented derive from the controlled synthetic benchmark and therefore do not constitute evidence of clinical prediction capability; definitive evaluation requires the real-data pipeline described in Section 2.5. The Stage-2 RL component failed to converge on the benchmark, and its viability for clinical action recommendation remains undemonstrated. The framework uses data from a single medical center (Beth Israel Deaconess Medical Center), and multi-center validation is necessary to assess generalizability across different patient populations, clinical practices, and documentation styles. The current single-decision formulation for Stage-2 does not capture the sequential nature of clinical decision-making over extended hospitalization periods. The approximately 72.9 million trainable parameters may limit real-time deployment in resource-constrained clinical settings. Finally, prospective clinical trials comparing AI-assisted versus standard decision-making would be necessary to establish clinical utility beyond retrospective performance metrics.

### 4.5. Future Work

The following priorities guide continued development: (1) implementing the real-data extraction pipeline from MIMIC-IV and conducting definitive evaluation with verified clinical outcomes, verified temporal separation, and per-task training; (2) establishing baseline comparisons against single-modality models (EHR-only, notes-only, imaging-only) and traditional clinical scoring systems (APACHE-III, SOFA) to quantify the marginal contribution of multimodal fusion; (3) extending Stage-2 from single-decision to multi-step reinforcement learning to capture treatment trajectories; (4) incorporating Bayesian uncertainty quantification through Monte Carlo dropout [43] or ensemble methods to provide calibrated confidence estimates for both predictions and action recommendations; (5) exploring federated learning [44] for multi-institutional training while preserving patient data privacy.

## 5. Conclusions

We presented a two-stage explainable multimodal architecture for post-surgical clinical decision support that integrates transformer-based multimodal fusion with Conservative Q-Learning. The framework addresses three critical gaps in current clinical AI: the gap between prediction and actionable decision support, the gap between black-box models and clinically interpretable systems, and—most fundamentally—the gap between inflated metrics from flawed data pipelines and genuine clinical prediction evidence.

We identified and formally characterized the target leakage problem in synthetic clinical data pipelines, where shared risk-score intermediaries create circular dependencies between inputs and targets that render performance metrics scientifically invalid. We proposed a rigorous real-data evaluation methodology using MIMIC-IV with verified clinical outcomes, event-censored temporal separation, uncertainty-weighted per-task training, and convergence-gated reinforcement learning deployment.

Controlled architectural validation confirmed component-level function: the encoder-fusion pipeline recovers embedded signals, the cross-modal attention mechanism learns differential modality weighting, and the explainability dashboard produces coherent visualizations. The Stage-2 reinforcement learning component did not converge on the benchmark, requiring investigation with real clinical data.

We believe that transparent identification of what works, what does not, and why—rather than selective presentation of favorable metrics—represents the most productive path toward trustworthy clinical AI systems. The architecture and evaluation methodology presented here provide the foundation for clinically valid multimodal decision support systems, pending implementation and evaluation on the proposed real-data pipeline.

## Data Availability

The MIMIC-IV, MIMIC-CXR, and MIMIC-IV-Note datasets are available from PhysioNet (https://physionet.org/) to credentialed researchers who complete the required human-subjects research training and sign the PhysioNet Credentialed Health Data Use Agreement. All source code used to implement the architecture, the controlled synthetic benchmark, and the scripts required to reproduce the experiments reported in this manuscript are publicly available at: https://github.com/mostakphoenixsoftbd/postsurgicalresearch/tree/master/Shareable/codes. No new patient data were generated by this study.

https://physionet.org/

## Ethics statement

This study used the MIMIC-IV, MIMIC-CXR, and MIMIC-IV-Note databases, which contain fully de-identified patient data made publicly available by the Massachusetts Institute of Technology Laboratory for Computational Physiology under the PhysioNet Credentialed Health Data Use Agreement. The original collection of these data was approved by the Institutional Review Boards of the Beth Israel Deaconess Medical Center and the Massachusetts Institute of Technology, with a waiver of informed consent granted because the data are de-identified. Because the present work involved only secondary analysis of these de-identified datasets, additional ethical approval was not required and individual patient consent could not be obtained or waived at the level of the present study.

## Notes

### Competing Interest Statement

The authors have declared no competing interest.

### Funding Statement

The author(s) received no specific funding for this work.

## References

1. Nepogodiev D, Martin J, Biccard B, Makupe A, Bhangu A. Global burden of postoperative death. Lancet. 2019;393(10170):401. doi:10.1016/S0140-6736(18)33139-8

2. Weiser TG, Regenbogen SE, Thompson KD, Haynes AB, Lipsitz SR, Berry WR, et al. An estimation of the global volume of surgery: a modelling strategy based on available data. Lancet. 2008;372(9633):139–144. doi:10.1016/S0140-6736(08)60878-8

3. Gawande AA, Thomas EJ, Zinner MJ, Brennan TA. The incidence and nature of surgical adverse events in Colorado and Utah in 1992. Surgery. 1999;126(1):66–75. doi:10.1067/msy.1999.98664

4. Rajkomar A, Oren E, Chen K, Dai AM, Hajaj N, Hardt M, et al. Scalable and accurate deep learning with electronic health records. npj Digit Med. 2018;1:18. doi:10.1038/s41746-018-0029-1

5. Shickel B, Tighe PJ, Bihorac A, Rashidi P. Deep EHR: a survey of recent advances in deep learning techniques for electronic health record (EHR) analysis. IEEE J Biomed Health Inform. 2018;22(5):1589–1604. doi:10.1109/JBHI.2017.2767063

6. Topol EJ. High-performance medicine: the convergence of human and artificial intelligence. Nat Med. 2019;25(1):44–56. doi:10.1038/s41591-018-0300-7

7. Holzinger A, Langs G, Denk H, Zatloukal K, Müller H. Causability and explainability of artificial intelligence in medicine. WIREs Data Min Knowl Discov. 2019;9(4):e1312. doi:10.1002/widm.1312

8. Tjoa E, Guan C. A survey on explainable artificial intelligence (XAI): toward medical XAI. IEEE Trans Neural Netw Learn Syst. 2021;32(11):4793–4813. doi:10.1109/TNNLS.2020.3027314

9. Ghassemi M, Oakden-Rayner L, Beam AL. The false hope of current approaches to explainable artificial intelligence in health care. Lancet Digit Health. 2021;3(11):e745–e750. doi:10.1016/S2589-7500(21)00208-9

10. Roberts M, Driggs D, Thorpe M, Gilbey J, Yeung M, Ursprung S, et al. Common pitfalls and recommendations for using machine learning to detect and prognosticate for COVID-19 using chest radiographs and CT scans. Nat Mach Intell. 2021;3(3):199–217. doi:10.1038/s42256-021-00307-0

11. Harutyunyan H, Khachatrian H, Kale DC, Ver Steeg G, Galstyan A. Multitask learning and benchmarking with clinical time series data. Sci Data. 2019;6:96. doi:10.1038/s41597-019-0103-9

12. Vaswani A, Shazeer N, Parmar N, Uszkoreit J, Jones L, Gomez AN, et al. Attention is all you need. In: Advances in Neural Information Processing Systems. Vol. 30. 2017. p. 5998–6008.

13. Li Y, Rao S, Solares JRA, Hassaine A, Ramakrishnan R, Canoy D, et al. BEHRT: transformer for electronic health records. Sci Rep. 2020;10:7155. doi:10.1038/s41598-020-62922-y

14. Rasmy L, Xiang Y, Xie Z, Tao C, Zhi D. Med-BERT: pre-trained contextualized embeddings on large-scale structured electronic health records for disease prediction. npj Digit Med. 2021;4:86. doi:10.1038/s41746-021-00455-y

15. Dosovitskiy A, Beyer L, Kolesnikov A, Weissenborn D, Zhai X, Unterthiner T, et al. An image is worth 16x16 words: transformers for image recognition at scale. In: International Conference on Learning Representations. 2021.

16. Hochreiter S, Schmidhuber J. Long short-term memory. Neural Comput. 1997;9(8):1735–1780. doi:10.1162/neco.1997.9.8.1735

17. Lipton ZC, Kale DC, Elkan C, Wetzel R. Learning to diagnose with LSTM recurrent neural networks. In: International Conference on Learning Representations. 2016.

18. Huang SC, Pareek A, Seyyedi S, Banerjee I, Lungren MP. Fusion of medical imaging and electronic health records using deep learning: a systematic review and implementation guidelines. npj Digit Med. 2020;3:136. doi:10.1038/s41746-020-00341-z

19. Acosta JN, Falcone GJ, Rajpurkar SA, Topol EJ. Multimodal biomedical AI. Nat Med. 2022;28(9):1773–1784. doi:10.1038/s41591-022-01981-2

20. Lu J, Batra D, Parikh D, Lee S. ViLBERT: pretraining task-agnostic visiolinguistic representations for vision-and-language tasks. In: Advances in Neural Information Processing Systems. 2019.

21. Komorowski M, Celi LA, Badawi O, Gordon AC, Faisal AA. The artificial intelligence clinician learns optimal treatment strategies for sepsis in intensive care. Nat Med. 2018;24(11):1716–1720. doi:10.1038/s41591-018-0213-5

22. Raghu A, Komorowski M, Celi LA, Szolovits P, Ghassemi M. Continuous state-space models for optimal sepsis treatment: a deep reinforcement learning approach. In: Machine Learning for Healthcare Conference. 2017.

23. Gottesman O, Johansson F, Komorowski M, Faisal A, Sontag D, Doshi-Velez F, et al. Guidelines for reinforcement learning in healthcare. Nat Med. 2019;25(1):16–18. doi:10.1038/s41591-018-0310-5

24. Kumar A, Zhou A, Tucker G, Levine S. Conservative Q-learning for offline reinforcement learning. In: Advances in Neural Information Processing Systems. Vol. 33. 2020. p. 1179–1191.

25. Levine S, Kumar A, Tucker G, Fu J. Offline reinforcement learning: tutorial, review, and perspectives on open problems. arXiv:2005.01643 [Preprint]. 2020. Available from: https://arxiv.org/abs/2005.01643

26. Lundberg SM, Lee SI. A unified approach to interpreting model predictions. In: Advances in Neural Information Processing Systems. Vol. 30. 2017. p. 4765–4774.

27. Sundararajan M, Taly A, Yan Q. Axiomatic attribution for deep networks. In: Proceedings of the 34th International Conference on Machine Learning. Vol. 70. 2017. p. 3319–3328.

28. Amann J, Blasimme A, Vayena E, Frey D, Madai VI. Explainability for artificial intelligence in healthcare: a multidisciplinary perspective. BMC Med Inform Decis Mak. 2020;20:310. doi:10.1186/s12911-020-01332-6

29. Wiens J, Saria S, Sendak M, Ghassemi M, Liu VX, Doshi-Velez F, et al. Do no harm: a roadmap for responsible machine learning for health care. Nat Med. 2019;25(9):1337–1340. doi:10.1038/s41591-019-0548-6

30. Johnson AEW, Pollard TJ, Berkowitz SJ, Greenbaum NR, Lungren MP, Deng CY, et al. MIMIC-CXR, a de-identified publicly available database of chest radiographs with free-text reports. Sci Data. 2019;6:317. doi:10.1038/s41597-019-0322-0

31. Johnson A, Pollard T, Horng S, Celi LA, Mark R. MIMIC-IV-Note: deidentified free-text clinical notes (version 2.2). PhysioNet. 2023. doi:10.13026/1n74-ne17

32. Alsentzer E, Murphy J, Boag W, Weng WH, Jindi D, Naumann T, et al. Publicly available clinical BERT embeddings. In: Proceedings of the 2nd Clinical Natural Language Processing Workshop. 2019. p. 72–78. doi:10.18653/v1/W19-1909

33. Hendrycks D, Gimpel K. Gaussian error linear units (GELUs). arXiv:1606.08415 [Preprint]. 2016. Available from: https://arxiv.org/abs/1606.08415

34. Lin TY, Goyal P, Girshick R, He K, Dollár P. Focal loss for dense object detection. In: Proceedings of the IEEE International Conference on Computer Vision. 2017. p. 2980–2988.

35. Kendall A, Gal Y, Cipolla R. Multi-task learning using uncertainty to weigh losses for scene geometry and semantics. In: Proceedings of the IEEE Conference on Computer Vision and Pattern Recognition. 2018. p. 7482–7491.

36. Johnson AEW, Bulgarelli L, Shen L, Gayles A, Shammout A, Horng S, et al. MIMIC-IV, a freely accessible electronic health record dataset. Sci Data. 2023;10:1. doi:10.1038/s41597-022-01899-x

37. Pang K, Li L, Ouyang W, Liu X, Tang Y. Establishment of ICU mortality risk prediction models with machine learning algorithm using MIMIC-IV database. Diagnostics. 2022;12(5):1068. doi:10.3390/diagnostics12051068

38. Pakbin A, Nowroozilarki Z, Lee DKK, Mortazavi BJ. Real-time mortality prediction using MIMIC-IV ICU data via boosted nonparametric hazards. arXiv:2110.08949 [Preprint]. 2021. Available from: https://arxiv.org/abs/2110.08949

39. Che Z, Purushotham S, Cho K, Sontag D, Liu Y. Recurrent neural networks for multivariate time series with missing values. Sci Rep. 2018;8:6085. doi:10.1038/s41598-018-24271-9

40. Irvin J, Rajpurkar P, Ko M, Yu Y, Ciurea-Ilcus S, Chute C, et al. CheXpert: a large chest radiograph dataset with uncertainty labels and expert comparison. In: Proceedings of the AAAI Conference on Artificial Intelligence. 2019.

41. Loshchilov I, Hutter F. Decoupled weight decay regularization. In: International Conference on Learning Representations. 2019.

42. Mamatov N, Kellmeyer P. Enhancing mortality prediction in cardiac arrest ICU patients through meta-modeling of structured clinical data from MIMIC-IV. arXiv:2510.18103 [Preprint]. 2025. Available from: https://arxiv.org/abs/2510.18103

43. Gal Y, Ghahramani Z. Dropout as a Bayesian approximation: representing model uncertainty in deep learning. In: Proceedings of the 33rd International Conference on Machine Learning. 2016.

44. Li T, Sahu AK, Talwalkar A, Smith V. Federated learning: challenges, methods, and future directions. IEEE Signal Process Mag. 2020;37(3):50–60. doi:10.1109/MSP.2020.2975749

